# EXTERNAL VALIDATION OF THE IMPROVING PARTIAL RISK ADJUSTMENT IN SURGERY (PRAIS2) MODEL FOR 30-DAY MORTALITY AFTER PEDIATRIC CARDIAC SURGERY

**DOI:** 10.1101/2020.04.16.20057513

**Authors:** Lucia Cocomello, Massimo Caputo, Rosie Cornish, Deborah A. Lawlor

**Author notes:** **Corresponding Author:** Lucia Cocomello, MRC Integrative Epidemiology Unit, Oakfield House, Oakfield Grove, Bristol, BS8 2BN, United Kingdom.

## Abstract

**Objective:** Risk stratification in paediatric patients undergoing heart surgery remains a challenge. The improving partial risk adjustment in surgery (PRAIS2) is a risk model predicting 30-day mortality which has been recently developed and validated using a UK-based cohort from April 2009-March 2015. We aimed to perform an independent temporal external validation to explore its generalisability and clinical utility.

**Methods:** PRAIS2 validation was carried out using a single centre (Bristol, UK) cohort from April 2004 to March 2009 and April 2015 to July 2019. For each subject PRAIS2 score was calculated according to the original formula. PRAIS2 performance was assessed in terms of discrimination by means of ROC curve analysis and calibration by using the calibration belt method.

**Results:** A total of 1330 (2004-2009) and 1187 (2015-2019) paediatric cardiac surgical procedures were included in the first and second independent validation, respectively (median age at the procedure 6.0 and 6.9 months). PRAIS2 score showed excellent discrimination for both independent validations (AUC 0.72 (95%CI: 0.65 to 0.80) and 0.87 (95%CI: 0.82 to 0.93), respectively). While PRAIS2 was only marginally calibrated in the first validation, with a tendency to underestimate risk P-value = 0.051), the second validation showed good calibration with 95% confidence belt containing the bisector for predicted mortality (P-value = 0.15); We also observed good performance in the subgroup of patients undergoing non-elective procedures (N = 482; AUC 0.78 (95%CI 0.68 to 0.87); Calibration belt containing the bisector (P-value=0.61).

**Conclusions:** In a single centre UK-based cohort, PRAIS2 showed excellent discrimination and calibration in predicting 30-day mortality in paediatric cardiac surgery including in those undergoing non-elective procedures. Our results support a wider adoption of PRAIS2 score in the clinical practice.

**Strengths and limitations of this study:**

- A strength of the present study is that data were prospectively collected as part of the UK National Congenital Heart Disease Audit and as such they undergo continuous and inclusive systematic validation that includes the review of a sample of case notes by external auditors to ensure coding accuracy.
- We used a recently proposed method (calibration belt) which does not require patients to be categorised according to risk percentile but rather provides a risk function across all risk value with relative uncertainty measure (95% CI)
- A key limitation of this study is that the sample size is relatively small and considerably smaller than the cohort used to develop PRAIS2

**Key questions:**

- **What is already known about this subject?** The improving partial risk adjustment in surgery (PRAIS2) is a risk model predicting 30-day mortality which has been recently developed and validated using a UK-wide cohort.
- **What does this study add?** The present study reported the first independent external validation of the PRAIS2 using a single centre cohort which confirmed excellent performance of the model and for the first time showed that it also accurately predicts mortality in patients undergoing non-elective procedures
- **How might this impact on clinical practice?** Our results support a wider adoption of the PRAIS2 in the clinical practice.

## INTRODUCTION

Congenital heart diseases (CHD) are the most common birth defect affecting between 6-8 per 1000 of live born children in middle and high income countries.^1 2^ Around 5000 paediatric cardiac surgical procedures are performed each year in the United Kingdom. Despite overall mortality being low (3%), there is notable variation in mortality rates between centres. Mortality rates, and variation between centres, are monitored by public and healthcare regulatory bodies as part of assessing the quality of care provided by the centres. In order to monitor a centre’s quality of care and performance and make fair comparisons between centres in terms of mortality following paediatric cardiac surgery it is essential to take account of differences in case mix across centres.^3^ This requires accurate risk stratification. However, CHD includes a large spectrum of diagnoses with a wide range of surgical procedures performed in the context of a relatively small number of patients. These characteristics make risk stratification extremely challenging.

Several methods that aim to provide an objective risk assessment for taking account of case mix when assessing centre performance have been developed. These include consensus based methods, such as the risk adjusted classification for congenital heart surgery (RACHS-1) ^4^ and Aristotle,^5^ and more recently empirical research based methods, such as the society of thoracic surgeons-European association of cardiothoracic surgery score (STS-EACTS)^6^ and the partial risk adjustment in surgery (PRAIS)^7^, which has been developed in the UK and proposed for measuring between centre variation in mortality across the UK. The most recently updated version, PRAIS2^8^, was developed to predict 30-day mortality using data from the UK National Congenital Heart Disease Audit (NCHDA).^9^ In comparison to PRAIS, PRAIS2 included more detailed information about acuity, diagnosis and comorbidities and was shown to have better discrimination and calibration than the original PRAIS^10^. However, adoption of PRAIS2 into clinical practice is still limited and this has been partially attributed to the lack of external independent validation other than the one presented by the authors in the original paper.11That validation used data from the same UK dataset but was undertaken on a temporally independent sub-cohort (Figure 1). Whilst such validation within a study is important, using the same cohort (and same group of investigators) can result in risk of optimism bias, which would tend to exaggerate risk stratification accuracy in comparison to findings from independent cohorts and investigators.^12,13^ Moreover, prediction models can present significant calibration drift due to temporal and geographic differences in case mix, patients characteristics and surgical technique; the impact of this on the performance of PRAIS2 is unknown.^14 15^ Lastly, as the data used to develop PRAIS2 did not include information on whether the procedures were elective or conducted as non-elective (emergencies, urgency and salvage) it was not possible to determine whether performance was similar in both of these situations. It is not implausible that stratification accuracy will differ between the two.

**Figure 1.**
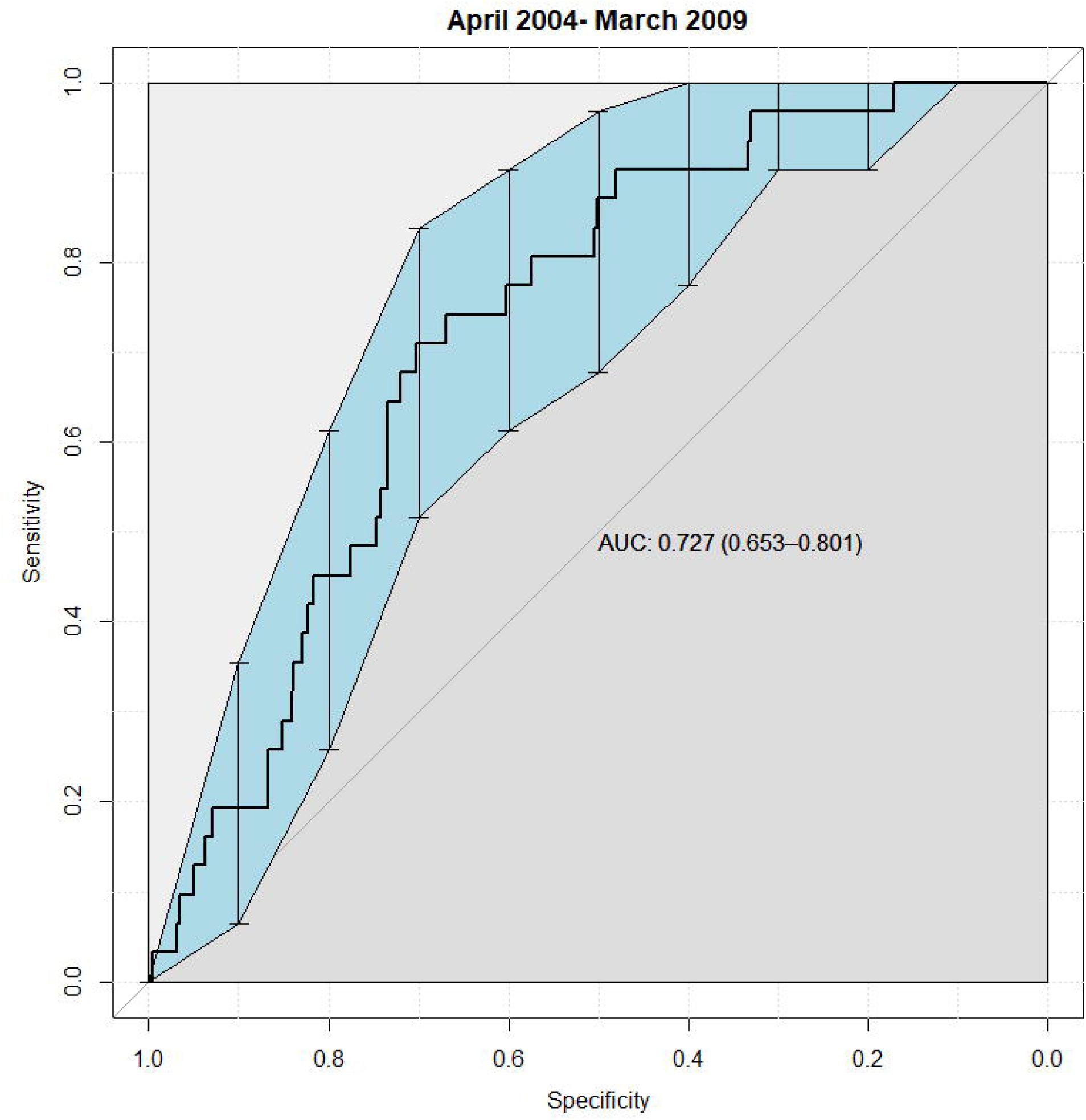
Patients included in the study.

The purpose of this study was to perform an external independent validation of the PRAIS2 score in a cohort from a single tertiary paediatric UK centre (in Bristol, South West England). This single UK centre contributed to the cohort in which PRAIS2 was originally developed but with data from different time periods than used here, where we explore performance separately in two cohorts: (i) undergoing procedures earlier than those used in PRAIS2 and (ii) undergoing procedures after those used in PRAIS2. To explore possible calibration drift, we have compared calibration of the model prediction between the three Bristol Heart Institute cohorts; procedures undertaken 2004-2009, 2009-2015 (included in PRAIS2 development) and 2015 to 2019. In a subsample, we were also able to undertake the first (exploratory) analysis of how well PRAIS2 performs in those undergoing non-elective procedures.

## METHODS

The study complies with the Declaration of Helsinki. This study was approved by the institutional review board (IRB) at the Bristol Heart Institute (reference number 250868). As this analysis came under clinical audit / quality of care assessment and all data were anonymised following the governance criteria of the NHS, the IRB agreed informed consent was not required.

### Patient and public involvement

This research was done without Patient and Public Involvement.

### Data Source

Data were obtained from the Bristol dataset which is part of the National Congenital Heart Disease Audit (NCHDA) within the National Institute of Cardiovascular Outcomes Research (NICOR). As such they undergo continuous and inclusive systematic validation of the data used here, which includes the review of a sample of case notes by external auditors to ensure coding accuracy.^9^ **Figure 1** summarises the data sources used in this study and their relationship to the UK National data used in the development of PRAIS2. The Bristol cohort includes data from patients undergoing procedures between April 2004 and July 2019. PRAIS2 was developed using national data (including that from the Bristol cohort) from patients undergoing procedures between April 2009 and March 2015. In these analyses we use Bristol data from April 2004 to March 2009 (N = 1330) and April 2015 to July 2019 (N = 1187) as independent external validation (hereafter referred to as independent validation 1 and 2, respectively). Whilst we acknowledge that treatments and mortality rates have changed since 2004, and the first validation set may not reflect contemporary practice, we would expect this to primarily affect model calibration; finding good discrimination for this earlier set would support external validity and generalisability of PRAIS-2. Seeing good discrimination across all time periods would provide valuable evidence about the generalisability of the PRAIS-2 model. In addition, we explored discrimination and calibration in the Bristol cohort that was also included in the national cohort to develop PRAIS2, April 2009 to March 2015 (N = 1794). This allows us to explore any evidence of geographical (centre) difference in stratification performance and using all three cohorts to explore the extent of temporal calibration drift.

The Bristol dataset included 4,885 paediatric cardiac surgical procedures (defined as surgery on the heart or great vessel in patients aged < 16 years old, excluding catheter procedures and trivial/minor procedures). Of these 4,885 we had to exclude 575 (12%) for missing data; the remaining 4310 contributed to one of the three temporal cohorts used here.

#### PRAIS2 prediction score and its constituent variables

PRAIS2 score is generated from a transformed logistic regression model of 30-day mortality following cardiac surgery.^10^ The model included the following perioperative variables: age, weight, diagnosis, procedure group, type of procedure, whether or not there was definite univentricular heart function, additional cardiac risk factors, acquired comorbidity, congenital comorbidity, severity of illness and an additional coefficient for procedures performed after 2013 (Supplementary Table 1 shows the units or categories of each of these variables). The formula for the PRAIS2 score (using the function from the logistic regression model) is

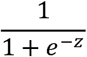

Where z is the logistic model function of the nine variables. In all analyses we used the PRAIS2 model (equation) that was recalibrated by the original authors after their internal validation (see Supplementary Equation 1).

#### Outcome

The primary outcome was 30-day all-cause mortality. Information on mortality was obtained from the Office for National Statistics (ONS).

### Statistical analysis

Continuous data were presented as medians with interquartile ranges (IQR). Categorical variables were presented as counts with percentages. We assessed the discriminative ability of the PRIAS2 model to distinguish between children who did and did not die within 30-days using the area under the receiver operating characteristic curve (AUC) analysis^16^ and its calibration by comparing observed to predicted 30-day mortality rates for groups of patients with different predicted levels of risk.^11 17 18^ Overall model calibration was assessed using the calibration belt method. The method is based on a generalisation of Cox’s regression modelling where the relation between the logits of the probability predicted by a model and of the event rates observed in a sample is represented by a polynomial function whose coefficients are fitted and its degree is fixed by a series of likelihood-ratio tests.^19^ Risk underestimation is suggested if the calibration belt with its 95% confidence interval (CI) is above the bisector (perfect prediction line) while overestimation is indicated by the belt with its 95% CI being below the bisector. The plot was accompanied by the Hosmer-Lemeshow test goodness of fit test statistical test.^20^ All analyses were performed using R version 3.5.2.

## RESULTS

Our analyses included 1125 patients undergoing 1330 procedures and 902 patients undergoing 1187 procedures in the external validation samples 1 and 2, respectively, and a subset (from validation 2) of 281, undergoing 482 non-elective procedures. Median age at the time of the first procedure of any participant included in any analyses was 6.5 months (IQR 1.6 – 42.5).

Table 1 shows patient characteristics of the two external validation cohorts and the subgroup undergoing non-elective procedures together with the Bristol cohort that was included in the national cohort used to develop PRAIS2.

**Table 1.** Patient characteristics in the earlier, external validation cohorts and in the non-elective subset. Definitions of variables are given in Supplementary Table 1

|  | **Independent validation 1**<br>**(April 2004 - March2009)** | **Independent validation 2**<br>**(April 2015 - July 2019)** | **Non-elective subset**<br>**(April 2015 - July 2019)** | **Cohort overlapping with**<br>**PRAIS2 cohort**<br>**(April 2009- March 2015)** |
| --- | --- | --- | --- | --- |
| **Total** | (n=1330 procedures<br>n=1125 patients) | (n=1187 procedures<br>n=902 patients) | (n=482 procedures<br>n=281 patients) | (n=1794 procedures<br>N=1461 patients) |
| **Age at procedure median,**<br>**months [IQR]** | 6.0 [1.4 - 27.7] | 6.9 [2.2-49.9] | 4.8 [1.1-35.5] | 6.0 [1.4-36.4] |
| **Weight median, Kg [IQR]** | 6.0 [3.4 – 12.0] | 6.9 [3.9-15.3] | 3.7 [3.1-5.3] | 6 [3.6-12.6] |
| **Diagnoses group n (%)<sup>a</sup>** |  |  |  |  |
| GROUP1 | 69 (5.1) | 107 (9.0) | 47 (9.8) | 153 (8.5) |
| GROUP2 | 149 (11.2) | 161 (13.6) | 77 (16.0) | 192 (10.7) |
| GROUP 3 | 109 (8.2) | 103 (8.7) | 65 (13.5) | 124 (9.3) |
| GROUP 4 | 215 (16.2) | 232 (19.5) | 118 (24.5) | 222 (12.4) |
| GROUP 5 | 104 (7.8) | 122 (10.3) | 49 (10.2) | 119 (6.6) |
| GROUP 6 | 98 (7.4) | 131 (11.0) | 44 (9.1) | 89 (4.9) |
| GROUP 7 | 175 (13.1) | 127 (10.7) | 34 (7.1) | 309 (17.2) |
| GROUP 8 | 150 (11.3) | 103 (8.7) | 32 (6.6) | 186 (10.4) |
| GROUP 9 | 20 (1.5) | <5 (<1) | <5 (<1) | 25 (1.4) |
| GROUP 10 | 56 (4.2) | 14 (1.2) | 6 (1.2) | 94 (5.2) |
| GROUP 11 | 185 (13.9) | 85 (7.2) | 8 (1.7) | 281 (15.6) |

| **Procedure Group n (%)<sup>a</sup>** |  |  |  |  |
| --- | --- | --- | --- | --- |
| GROUP 1 | <5 (<1) | 31 (2.6) | 22 (4.6) | 27 (1.5) |
| GROUP 2 | 29 (2.2) | 11 (0.9) | 10 (2.1) | 33 (1.8) |
| GROUP 3 | 105 (7.9) | 46 (3.9) | 34 (7.1) | 84 (4.6) |
| GROUP 4 | 99 (7.4) | 103 (8.7) | 73 (15.1) | 114 (6.3) |
| GROUP 5 | 236 (17.8) | 228 (19.2) | 126 (26.1) | 344 (19.1) |
| GROUP 6 | 148 (11.1) | 53 (4.5) | 36 (7.5) | 179 (9.9) |
| GROUP 7 | 21 (1.6) | 44 (3.7) | 16 (3.3) | 49 (2.7) |
| GROUP 8 | 79 (5.9) | 96 (8.1) | 12 (2.5) | 103 (5.7) |
| GROUP 9 | 20 (1.5) | 16 (1.3) | 5 (1.0) | 26 (1.4) |
| GROUP 10 | 36 (2.7) | 35 (2.9) | 7 (1.5) | 70 (3.9) |
| GROUP 11 | 36 (2.7) | 19 (1.6) | 8 (1.7) | 57 (3.1) |
| GROUP 12 | 32 (2.4) | 63 (5.3) | <5 (<1) □ | 51 (2.8) |
| GROUP 13 | 80 (6.0) | 66 (5.6) | 5 (1.0) | 103 (5.7) |
| GROUP 14 | 38 (2.8) | 43 (3.6) | 7 (1.5) | 55 (3.0) |
| GROUP 15 | 216 (16.2) | 144 (12.1) | 11 (2.3) | 273 (13.8) |
| GROUP 20 | 154 (11.6) | 189 (15.9) | 109 (22.6) | 226 (12.5) |
| **Severity of illness n(%)** | 0 (0) | 221 (18.6) | 202 (41.9) | 86 (4.7) |
| **Acquired comorbidity n (%)** | 9 (0.7) | 205 (17.3) | 131 (27.2) | 94 (5.2) |
| **Congenital comorbidity n (%)** | 172 (12.9) | 341 (28.7) | 143 (29.7) | 268 (14.9) |
| **Additional cardiac risk factors n (%)** | 58 (4.4) | 81 (6.8) | 55 (11.4) | 52 (2.8) |
| **UVH category n (%)** | 141 (10.6) | 217 (18.3) | 95 (19.7) | 258 (14.3) |
| **Bypass n (%)** | 884 (66.4) | 902 (76.0) | 272 (56.4) | 1333 (74.3) |
| **PRAIS2 mean [SD]** | 2.1% [±2.8] | 2.7% [±4.0] | 5.2% [±4.2] | 2.2% [±3.3] |
| **30-day mortality** | 31 (2.3) | 23 (1.9) | 22 (4.6) | 56 (3.1) |

<sup>a</sup> as the diagnoses groups and procedure groups have lengthy text details, we have not provided them in the table; they can be found in Supplementary Table 1.
☐ Exact number suppressed for disclosure control purposes

There was a trend towards an increased risk profile (increasing in the number of complex procedures, group 1) in the most recent cohort and a subsequent increase in the expected mortality (PRAIS2). Notably, the observed mortality trend was in the opposite direction, with a reduction in the most recent cohort.

The predicted risk in the external validation cohorts 1 and 2, the PRAIS2 overlapping cohort and the non-elective subset were 2.1%,2.7%, 5.2% and 2.2% respectively and the observed mortality was 2.3%, 1.9%, 4.6% and 3.1%) respectively. Individual risk distribution among those who died and those who survived is shown in Supplementary Figure 1.

In the cohort 2 external validation analysis, the PRAIS2 score showed good discrimination ability (AUC 0.87; 95%CI 0.82-0.93, Figure 2b) and calibration between predicted and observed mortality across all risk categories with 95% confidence interval of calibration belt containing the bisector (p value=0.16,; Figure 3b;. Hosmer and Lemeshow goodness of fit test chi-squared=7.60, df=8, P-value 0.47, Supplementary Table 2b).

**Figure2.**
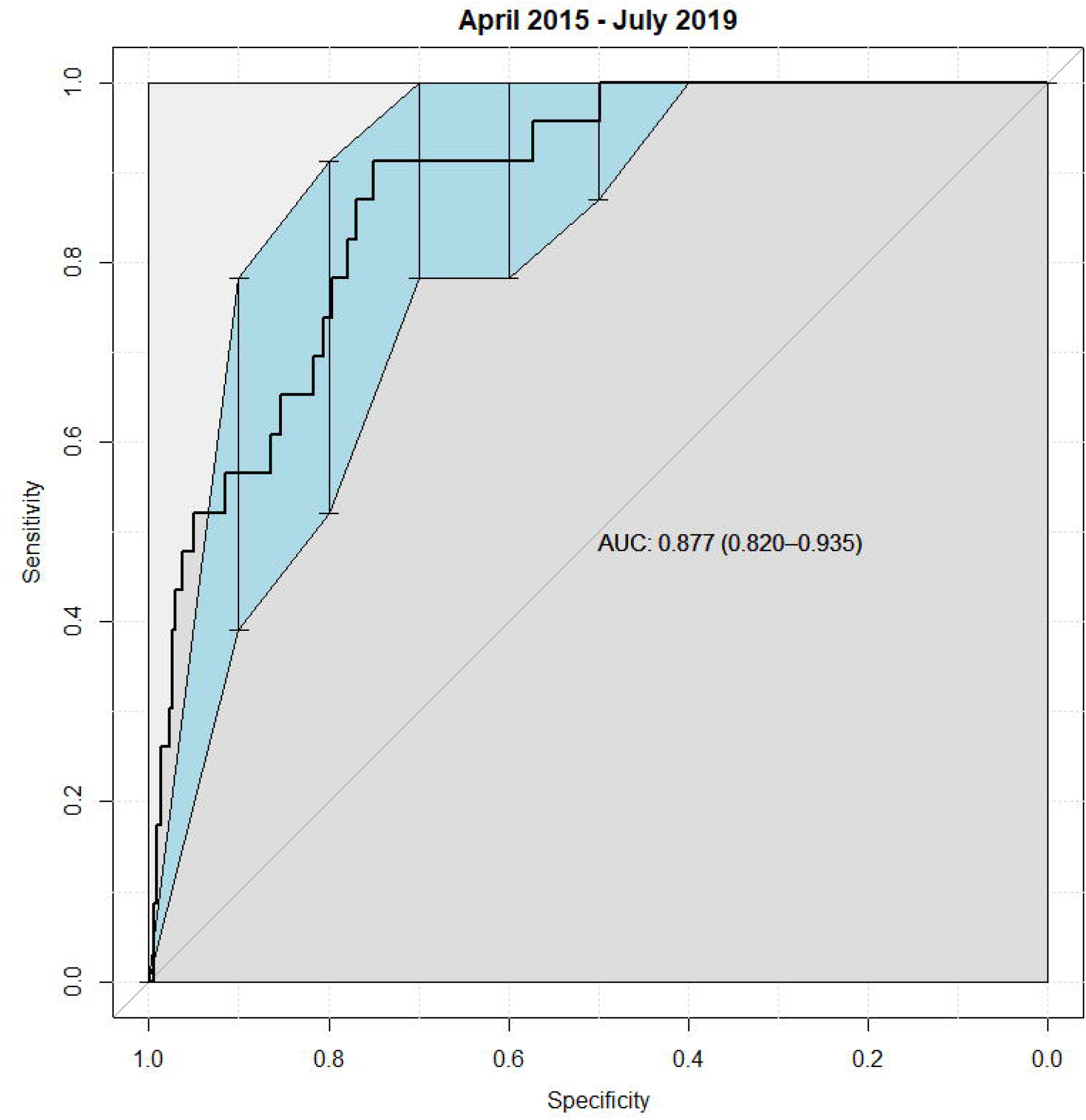
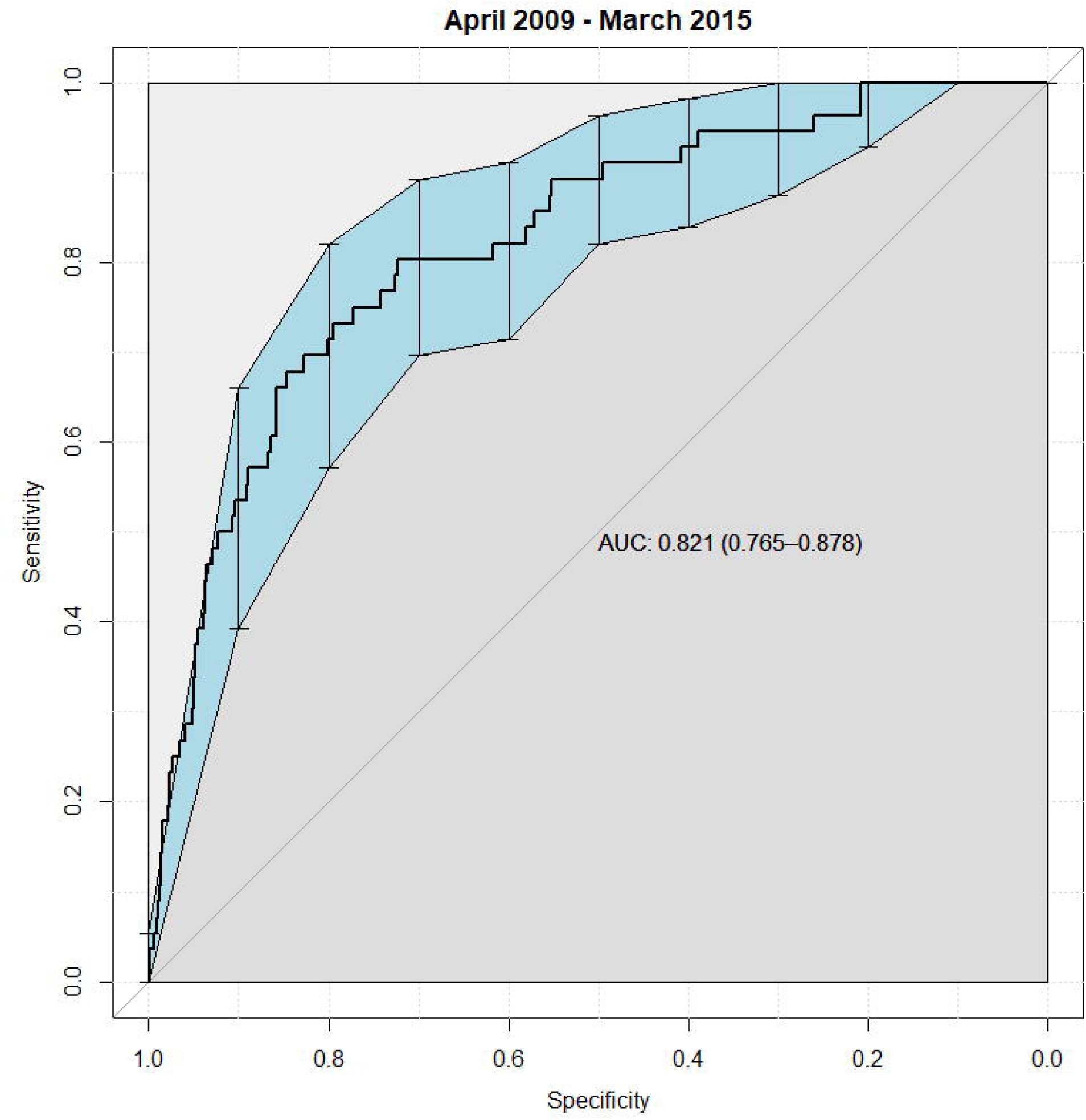
ROC curve for PRAIS2 in the independent validation 1 and 2 (2a and 2b) and overlapping (2c) cohorts.

**Figure3a.**
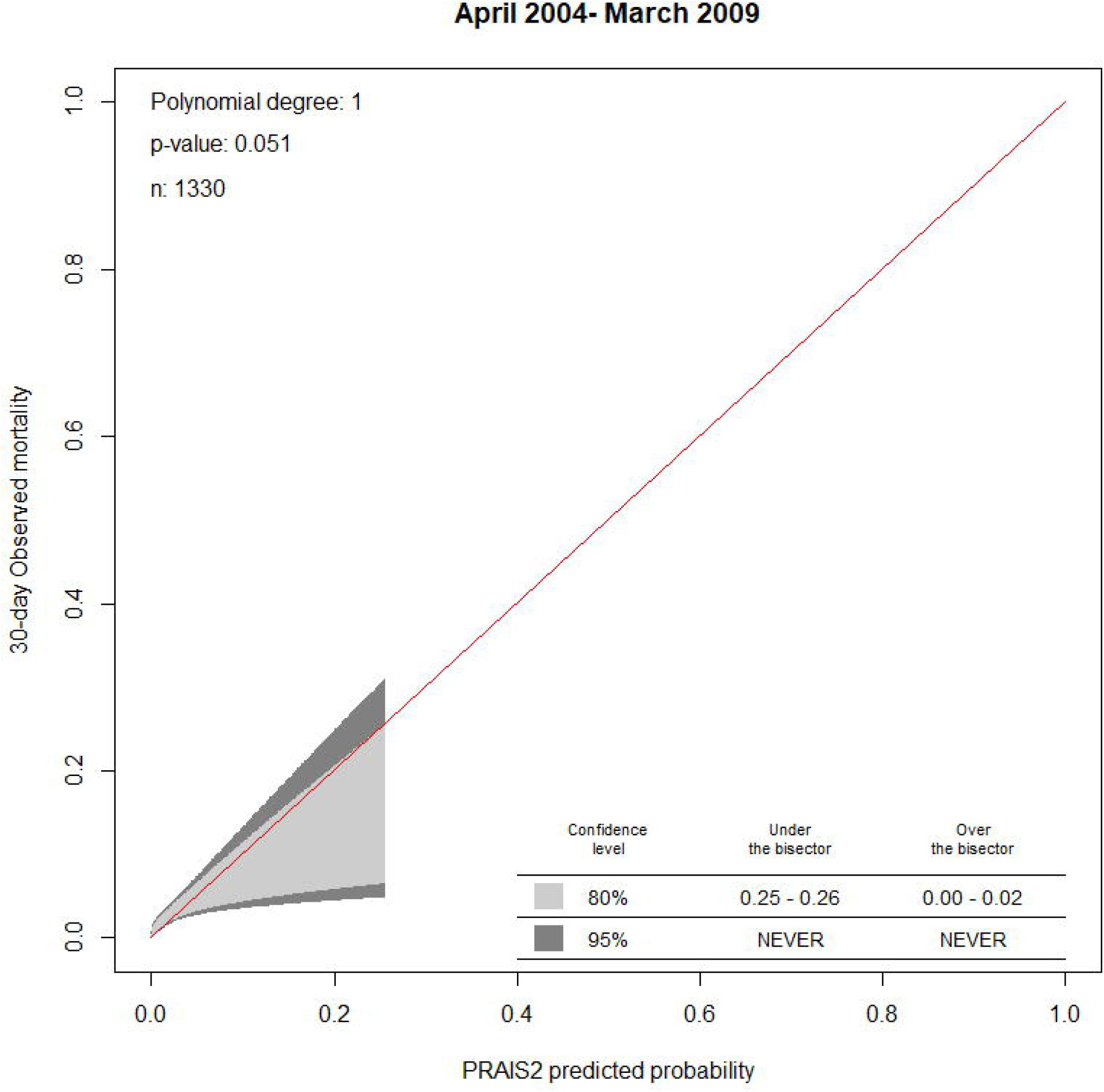
Calibration belt of PRAIS2 in the independent validation 1.

**Figure3b.**
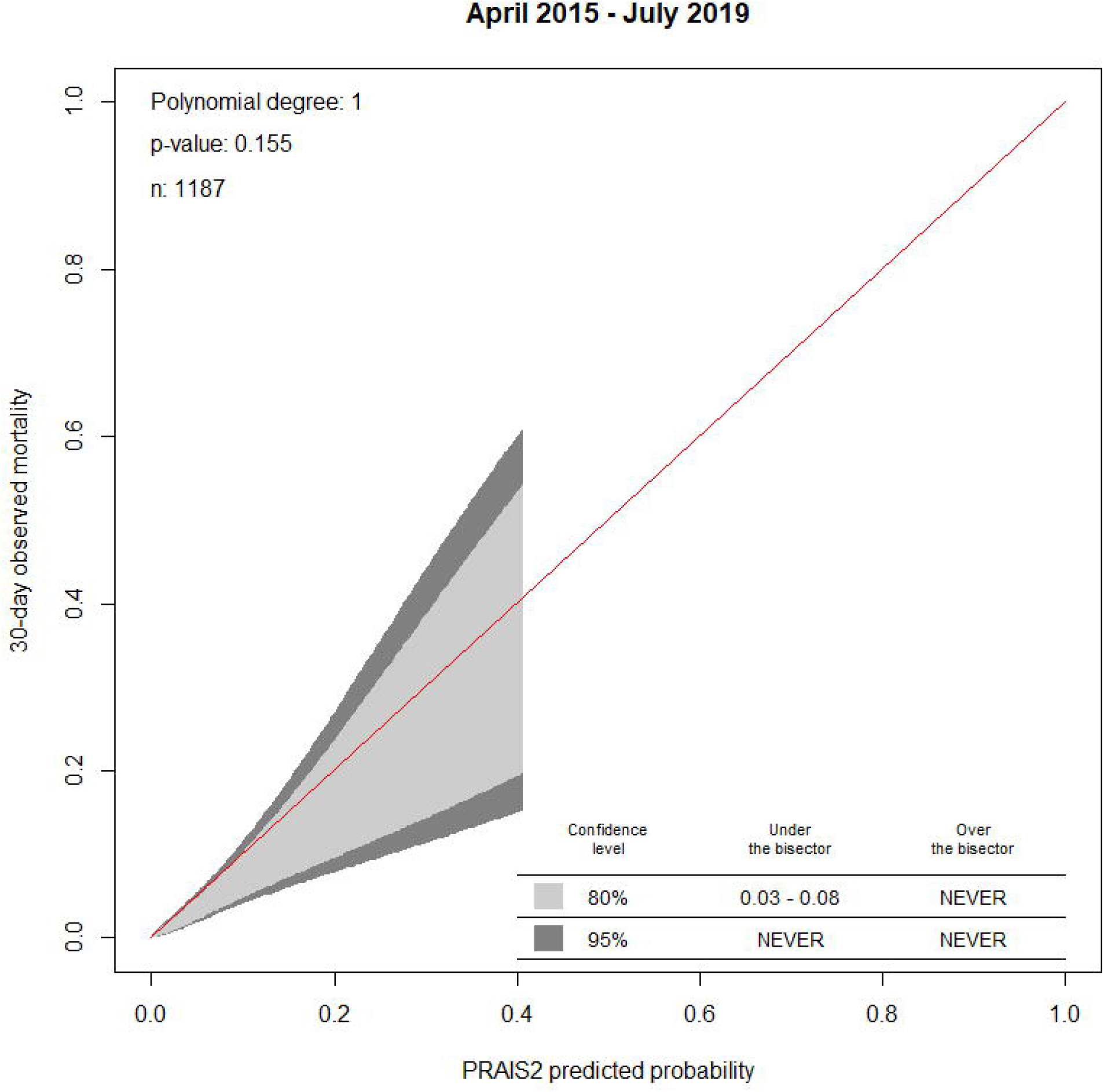
Calibration belt of PRAIS2 in the independent validation 2.

**Figure3c.**
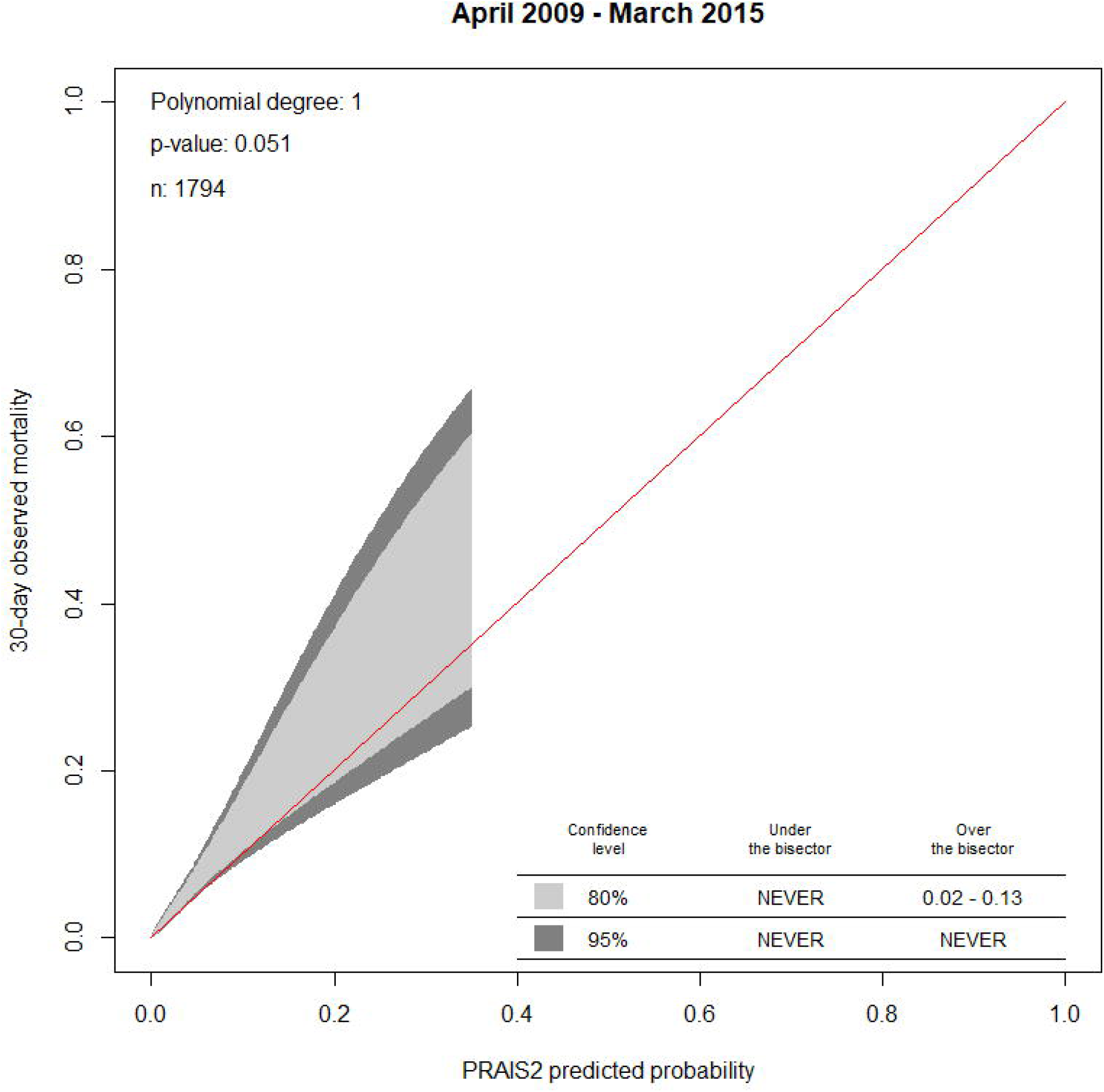
Calibration belt of PRAIS2 in the and overlapping cohorts.

In cohort 1, PRAIS2 showed a lower but still acceptable discrimination ability (AUC 0.72 (95%CI: 0.65 to 0.80, Figure 2a) and was only marginally calibrated according to the calibration belt method (p-value=0.051, Figure 3a; Hosmer and Lemeshow goodness of fit test chi-squared=15.80, df=8, P-value 0.045, Supplementary Table 2a).

In additional analyses to check that data from Bristol were not notably different to the main UK wide discovery cohort, we showed that the cohort from Bristol that contributed to the original PRAIS2 discovery also had good discrimination, with results similar to the whole PRAIS2 discovery cohort (AUC 0.82 (95%CI 0.77-0.88, Figure 2c). However, the model was only marginally calibrated with a tendency to risk underestimation (Figure 3c). Hosmer and Lemeshow goodness of fit test chi-squared =21.90; df=8; P-value=0.0051; Supplementary Table 2c). In the subgroup of patients undergoing non-elective procedures, the model showed good discrimination, with AUC 0.78 (95%CI 0.68-0.87, Figure 4a) and good calibration, with the bisector contained within the 95% confidence level of the calibration belt. (p-value 0.61; Figure 4b and Hosmer and Lemeshow goodness of fit test, chi-squared=10.04, df=8, P-value=0.26; Supplementary Table 2d). We were not able to conduct the analysis restricted to elective procedures as only one participant died in this subset.

**Figure 4a.**
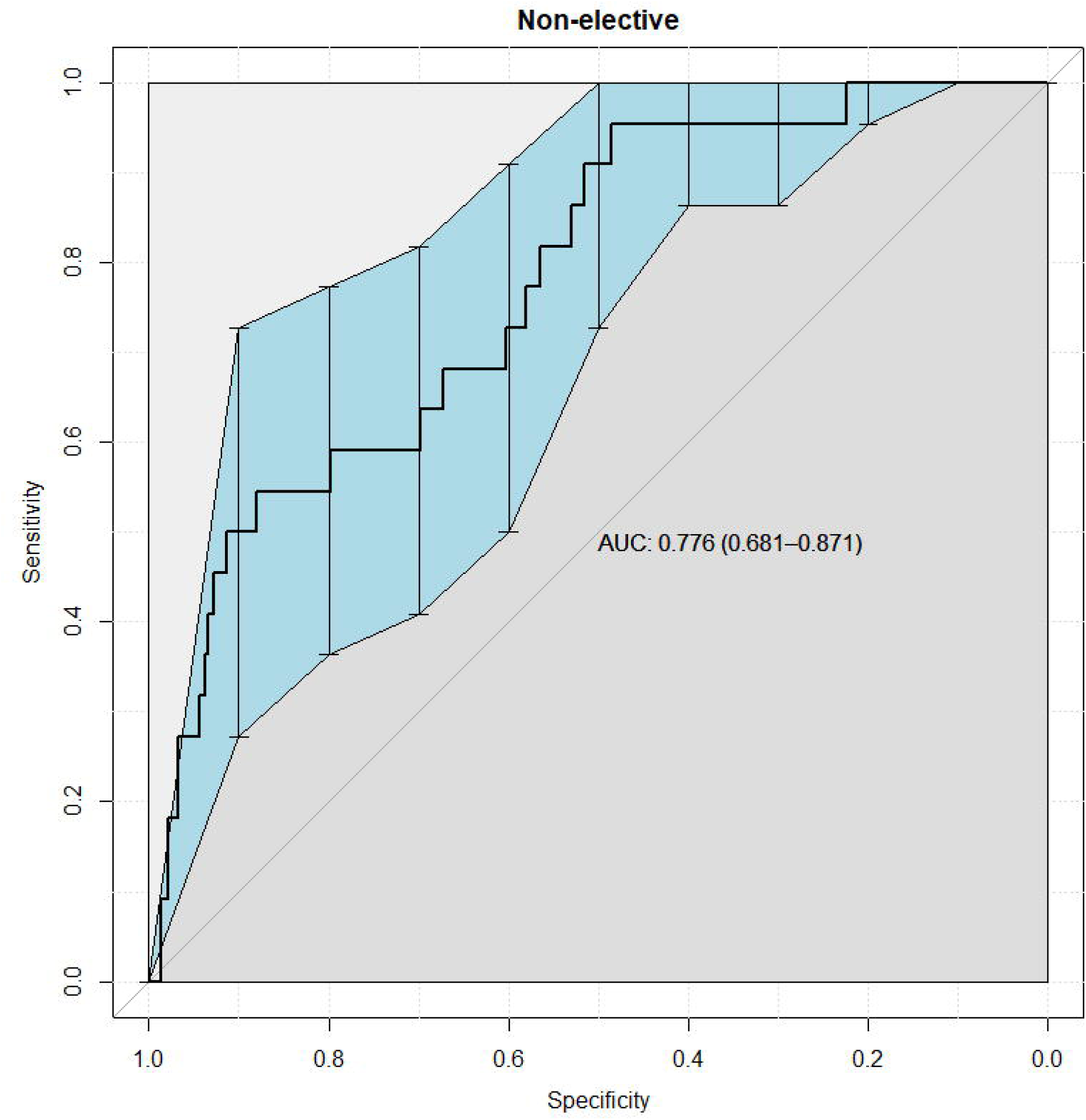
AUC in the subset of non-elective procedures in the external validation cohort.

**Figure 4b.**
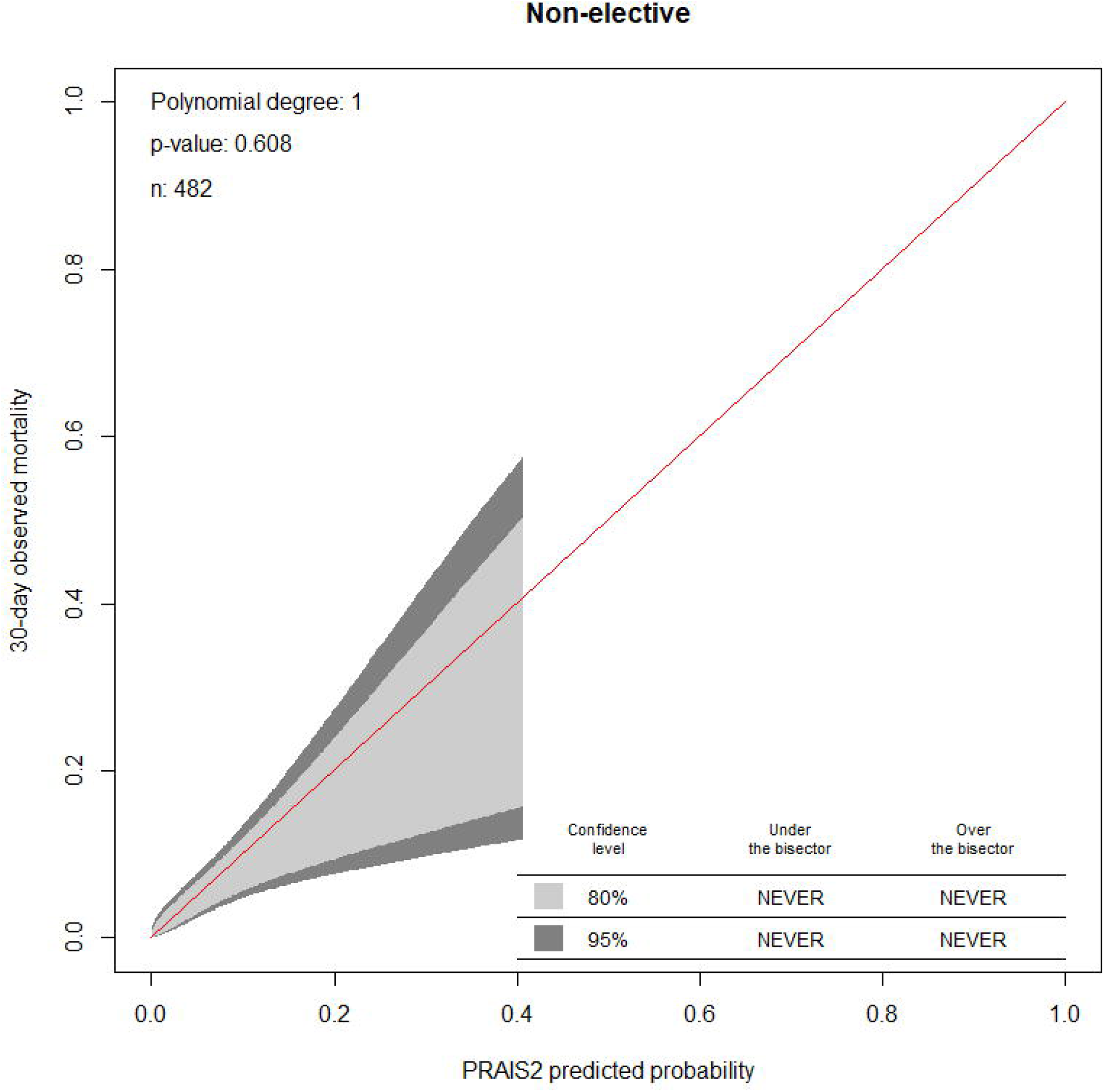
Calibration belt in the subset of non-elective procedures in the external validation cohort.

## DISCUSSION

Risk prediction models play an important role in current paediatric cardiac surgical practice, because they allow meaningful comparison of outcomes between institutions by adjusting for differing case-mix. Moreover, they can also give useful information for surgical decision making, preoperative patient information and quality assurance measures. Before implementing a risk model in clinical practice, it is important to confirm its prediction ability with external validation.^11 21^

To our knowledge PRAIS2 has never undergone external validation other than that originally presented by the authors who developed the model. It is important that prediction models are validated by independent researchers, as well as in independent samples, before being adopted into clinical practice because authors evaluating the performance of their own model may tend to be overly optimistic in interpreting results or selective reporting.^13 22^ For this reason we conducted an external validation study of PRAIS2 in two cohorts from the paediatric cardiac surgical procedures cohort at the Bristol Royal Hospital for Children. Our findings show that in these two, independent (of the original cohort used to develop PRAIS2), cohorts of 1330 procedures (in 1125 patients) and 1187 procedures (in 902 patients), respectively, the PRAIS2 model has good discrimination. Overall, the model showed good calibration. Remarkably, the model showed the best calibration in the most recent cohort. These findings do not suggest a relevant calibration drift. However, a further validation of the PRAIS2 in the coming years is recommended as calibration drift is plausible and can result in risk overestimation.^15^

As PRAIS2 was developed with data from across the UK and in our external validation we have only used data from Bristol we wanted to make sure that prediction in Bristol was broadly consistent with the UK as a whole. We therefore explored the predictive accuracy of PRAIS2 in the Bristol cohort that contributed to the original PRAIS2 discovery and found good discrimination, but the model was only marginally calibrated with a tendency to underestimate risk. It is not uncommon for calibration to differ between cohorts from different regions or over time. We are not able to determine variation in calibration between all of the different centres that contributed to the original PRAIS2 development as data were not presented by centre.^10^ It is interesting that a more recent Bristol cohort was better calibrated to PRAIS2 than the original Bristol cohort that contributed to PRAIS2 development. This could be partially explained by improved surgical results during recent years in our centre.

PRAIS2 development excluded non-elective procedures and so its accuracy in this group, or whether this would be a valuable predictor, is unknown. We demonstrated that PRAIS2 does have good discrimination and calibration in this higher risk subgroup. However, we acknowledge that our numbers were small for these analyses and this needs further exploration and validation in larger independent samples. We were not able to compare performance between those undergoing elective procedures and those undergoing non-elective procedures as there were insufficient deaths in those undergoing elective procedures. This marked difference in mortality between elective and non-elective procedures suggests that elective vs non-elective status might be a valuable covariable in PRAIS2, but large cohorts with these data would be needed to determine this.

A strength of the present study is that data were prospectively collected as part of the NCHDA national audit and as such they undergo continuous and inclusive systematic validation that includes the review of a sample of case notes by external auditors to ensure coding accuracy.^9^ A key limitation of this study is that the sample size is relatively small and considerably smaller than the cohort used to develop PRAIS2, which included a total of 21838 procedures (combining discovery and validation). Unlike the development cohort that included all UK patients, we only have data from one centre located in the South West of England where the population are more affluent and less ethnically mixed in comparison to the UK as a whole. Hence further replication in a larger and more diverse population is necessary.

The Hosmer and Lemeshow goodness of fit test for calibration is widely used in assessing calibration,^23 24^ but it provides a statistical test result comparing predicted to observed outcome rates across arbitrary grouping of deciles of risk, which may have limited clinical applicability. Therefore, we used a recently proposed method (calibration belt) which does not require patients to be categorised but rather compares across the continuous score of risk for our main method of assessing calibration.^25^ We also presented Hosmer and Lemeshow goodness of fit test in the supplementary material for comparison with any previous studies.

In conclusion, our study shows good external validity of PRAIS2 for predicting short-term mortality in paediatric cardiac surgery (the outcome it was developed to predict). We also show preliminary evidence that PRAIS2 accurately predicts short-term mortality in non-elective procedures. If similar results were found in more diverse external populations, including those outside of the UK, its use as the standard tool for risk prediction in paediatric patients undergoing cardiac surgery would be recommended.

## Data Availability

The authors confirm that the data supporting the findings of this study are available within the article or its supplementary materials.

## Acknowledgments

We thank Mai Baquedano for support with data management

---

**Supplementary Figure 1.**
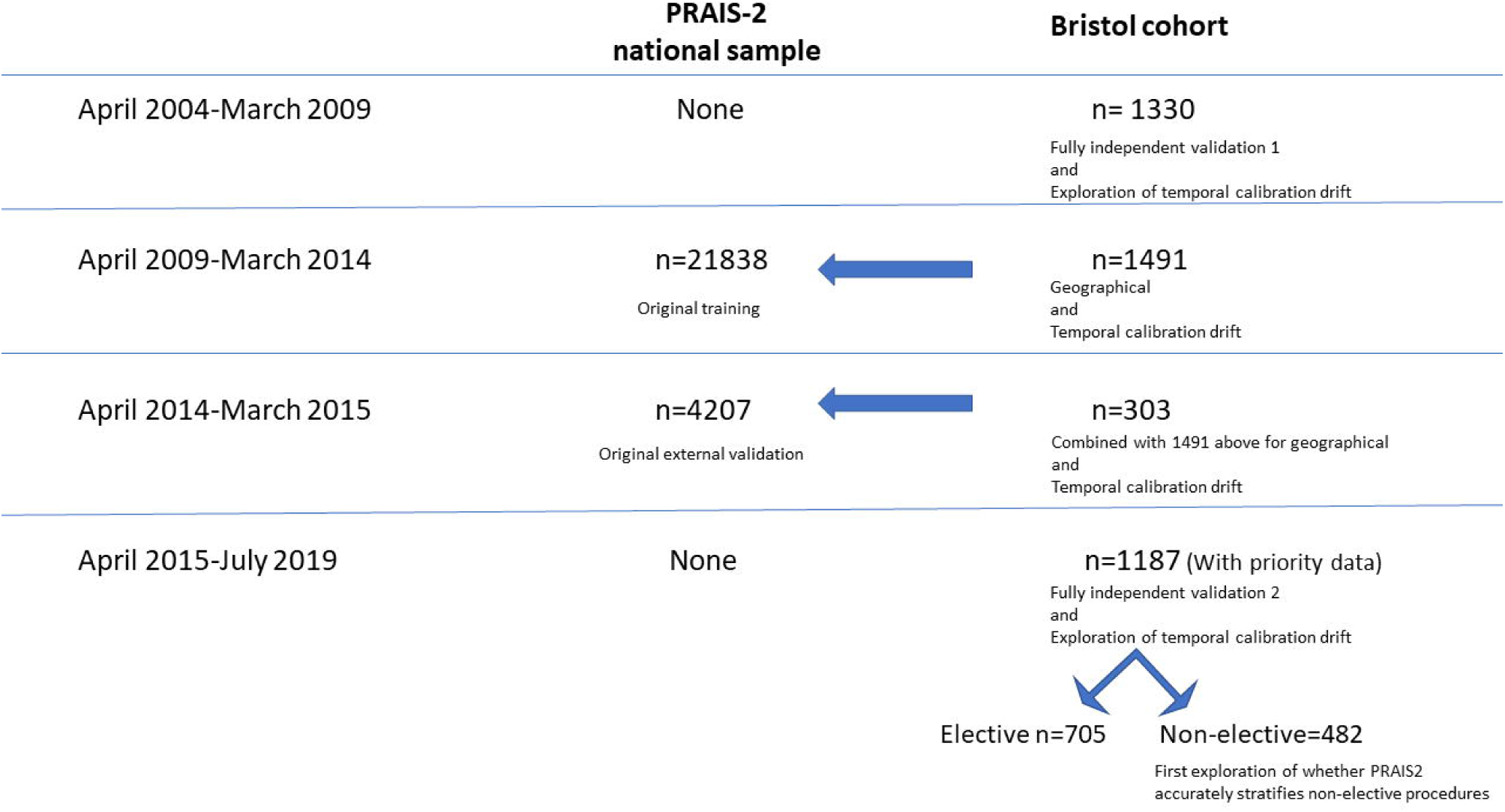
Sinaplot of PRAIS2 distribution stratified for incidence of 30-day mortality.

## Contributorship Statement

**Lucia Cocomello:** conception and design, data acquisition, analysis and interpretation of data, writing publication

**Massimo Caputo**: Critical revision of publication, approval of final publication

**Rosie Cornish:** Critical revision of publication, approval of final publication

**Deborah Lawlor:** Writing Publication, Critical revision of publication, approval of final publication, supervision

